# HIV-1 drug resistance in people on dolutegravir-based ART: Collaborative analysis of cohort studies

**DOI:** 10.1101/2023.04.05.23288183

**Authors:** Tom Loosli, Stefanie Hossmann, Suzanne M. Ingle, Hajra Okhai, Katharina Kusejko, Johannes Mouton, Pantxika Bellecave, Ard van Sighem, Melanie Stecher, Antonella d’Arminio Monforte, M. John Gill, Caroline A. Sabin, Gary Maartens, Huldrych F. Günthard, Jonathan A. C. Sterne, Richard Lessells, Matthias Egger, Roger Kouyos

## Abstract

**Background:** The widespread use of the integrase strand transfer inhibitor (INSTI) dolutegravir (DTG) in first- and second-line antiretroviral therapy (ART) may facilitate emerging resistance. We combined data from HIV cohorts to examine patterns of drug resistance mutations (DRMs) and identify risk factors for DTG resistance.

**Methods:** Eight cohorts from Canada, Europe, and South Africa contributed data on individuals with genotypic resistance testing on DTG-based ART. Resistance levels were categorised using the Stanford algorithm. We identified risk factors for resistance using mixed-effects ordinal logistic regression models.

**Results:** We included 750 people with genotypic resistance testing on DTG-based ART between 2013 and 2022. Most had HIV subtype B (N=444, 59·2%) and were treatment-experienced; 134 (17.9%) were on DTG dual and 19 (2.5%) on DTG monotherapy. INSTI DRMs were detected in 100 (13·3%) individuals; 21 (2·8%) had more than one mutation. Most (N=713, 95·1%) were susceptible to DTG, 8 (1·1%) had potential-low, 5 (0·7%) low, 18 (2·4%) intermediate and 6 (0·8%) high-level DTG resistance. The risk of DTG resistance was higher on DTG monotherapy (adjusted odds ratio (aOR) 37·25, 95% CI 11·17 to 124·2) and DTG lamivudine dual therapy (aOR 6·59, 95% CI 1·70 to 25·55) compared to combination ART, and higher in the presence of potential-low/low (aOR 4.62, 95% CI 1.24 to 17.2) or intermediate/high-level (aOR 7·01, 95% CI 2·52 to 19·48) nucleoside reverse transcriptase inhibitors (NRTI) resistance. Viral load on DTG showed a trend towards increased DTG resistance (aOR 1·42, 95% CI 0·92 to 2·19 per standard deviation of log_10_ area under the viral load curve).

**Interpretation:** Among people experiencing virological failure on DTG-based ART, INSTI DRMs were uncommon, and DTG resistance was rare. DTG monotherapy and NRTI resistance substantially increased the risk for DTG resistance, which is of concern, notably in resource-limited settings.

**Funding:** US National Institutes of Health, Swiss National Science Foundation.

**Research in context:** *Evidence before this study:* We searched SCOPUS on 20 March 2023 for all publications from inception using the terms “dolutegravir” or “DTG”, “resistant” or “resistance”, and “HIV”. The available evidence on resistance evolution in people living with HIV (PLHIV) with virological failure on DTG-based ART is limited. Most studies assessed the efficacy of DTG-based regimens in clinical studies and reported drug resistance in individuals experiencing virological failure as a secondary objective or reported single or multiple cases of patients developing resistance on DTG-based ART. Clinical trials such as the NADIA trial showed a high degree of viral suppression even in people with NRTI resistance. Consequently, previous analyses included only a small number of people experiencing failure on DTG; the SINGLE trial with 39 people with virologic failure on DTG was the largest. The highest number of individuals with DTG resistance was nine study participants in the NADIA trial. There is evidence that DTG resistance in PLHIV on a DTG monotherapy may be more likely. Other studies suggest that HIV subtype and mutations acquired during a first-generation INSTI-based regimen might affect the risk of DTG resistance.

*Added value of this study:* To our knowledge, this is the first study systematically investigating resistance in PLHIV experiencing virologic failure on DTG-based ART using a multi-cohort collaboration design reflecting real-world routine care. We collected genotypic resistance tests and clinical data from eight observational HIV cohorts. This resulted in a large dataset of PLHIV experiencing virologic failure on a DTG regimen (over 700 individuals). It allowed a robust assessment of drug resistance mutations and risk factors for DTG resistance. Cross-resistance of first-generation INSTIs does not appear to explain the mutation patterns in HIV-infected individuals who experience virological failure on DTG-based ART regimens. PLHIV who received DTG monotherapy or DTG lamivudine dual therapy and those infected with non-B subtypes were more likely to develop resistance. Resistance to NRTIs was a major risk factor for DTG resistance, indicating that PLHIV receiving functional monotherapy are more likely to develop DTG resistance.

*Implications of all the available evidence:* HIV drug resistance is a significant threat to the sustainability of current and future antiretroviral therapy for combating the ongoing HIV pandemic. Our collaborative analysis shows that cases of DTG resistance are so far rare but not negligible. Given the global DTG roll-out, this might lead to increased frequencies and transmission of DTG resistance, particularly in PLHIV with resistance to NRTIs. While the evidence regarding subtype differences is tentative, it indicates that non-B subtypes, which are most relevant for the global roll-out of DTG, might be associated with an increased risk of resistance.

## Introduction

The integrase strand transfer inhibitor (INSTI) dolutegravir (DTG) was approved in 2013 in the United States and shortly afterwards in the European Union to treat HIV infection. In 2019, the World Health Organization (WHO) recommended DTG as the preferred drug for first-line and second-line antiretroviral therapy (ART) in all populations, including pregnant women and those of childbearing age. Since then, DTG-based ART was rolled out globally,^1^ with about 100 countries including DTG in their treatment guidelines by mid 2020.^2^

DTG has a high genetic barrier to resistance,^3,4^ and relatively few people living with HIV (PLHIV) are so far known to have developed resistance.^5–7^ The mutations leading to DTG resistance may differ between HIV subtypes. In INSTI-naïve PLHIV, DTG resistance is mainly associated with the R263K mutation,^8,9^ which was observed in three cases of DTG resistance in the NADIA trial.^10^ The N155H mutation was present in two individuals with subtype A and C in the SAILING trial,^11^ while the G118R mutation appears to be facilitated by a natural polymorphism in subtype C.^12^ In a recent study in Ethiopia, the Q148H/K/R mutation was found to be less prevalent in subtype C.^13^ Pre-existing mutations, such as those acquired during a first-generation INSTI regimen, may directly confer resistance to DTG or affect the accumulation of additional mutations.^14,15^

The risk factors and the mutational patterns that confer resistance to DTG in vivo are less well established than for older antiretroviral drugs.^16^ The widespread use of DTG in resource-limited settings, where ART regimens are highly standardised, drugs are recycled, access to adherence support, viral load and resistance testing is limited, may facilitate the emergence of resistance. We combined data from European, North American, and South African cohorts to identify risk factors for DTG resistance and to examine the patterns of resistance mutations across different HIV subtypes.

## Methods

### Data sources

We pooled data from eight HIV cohorts, including six European, one Canadian, and one South African cohort: the AIDS Therapy Evaluation in the Netherlands cohort (ATHENA),^17^ the Agence Nationale de la Recherche sur le SIDA et les hépatites virales (ANRS CO3), Aquitaine Cohort,^18^ the Italian Cohort of Antiretroviral-Naïve Patients (ICONA),^19^ the Köln/Bonn Cohort (CBC), Germany,^20^ the UK Collaborative HIV Cohort (UK CHIC) Study (and linked UK HIV Drug Resistance Database (UKHDRD)),^21^ the Swiss HIV Cohort Study (SHCS),^22^ the South Alberta Clinic Cohort (SAC), Canada,^23^ and the South African Aid for AIDS (AfA) cohort.^24^ The European and North-American cohorts (apart from UK CHIC) participate in the ART Cohort Collaboration (ART-CC)^25^ and AfA in the International epidemiology Databases to Evaluate AIDS (IeDEA).^26^ The clinical data were provided by the data centres of the two cohort collaborations, and the genotypic data by the cohorts.

### Inclusion criteria

Participants who underwent genotypic resistance testing from plasma HIV-1 RNA covering the integrase gene between two weeks after starting and up to two months after stopping any DTG-based regimen were eligible. In the case of multiple genotypic resistance tests, the latest was considered. Participants with unknown dates of initiation of DTG-based ART were excluded. The analysis of risk factors for DTG resistance was restricted to individuals with at least one year of follow-up.

### HIV drug resistance

We defined two HIV drug resistance outcomes: the level of resistance to DTG and the presence of known drug resistance mutations (DRMs). The Stanford HIV Database version 9·0 and the Stanford HIVdb algorithm^27^ were used to categorise drug resistance levels as susceptible (score below 10), potential low (10-14), low (15-29), intermediate (30-59) or high (≥60). The same approach was used to assess resistance to all other antiretroviral drugs, whereby drug resistance to tenofovir alafenamide (not covered by the Stanford algorithm) was considered equal to tenofovir resistance. Resistance to non-nucleoside reverse transcriptase inhibitors (NNRTIs) was calculated as the median of the scores for efavirenz, etravirine, nevirapine, and rilpivirine. Resistance to nucleoside reverse transcriptase inbibitors (NRTIs) was calculated as the median of abacavir, zidovudine, emtricitabine/lamivudine, and tenofovir scores (see sensitivity analyses for alternative definitions).

### HIV subtyping

We determined HIV subtypes from the integrase gene using COMET (COntext-based Modeling for Expeditious Typing)^28^ and REGA.^29^ If REGA and COMET output differed, the subtype with higher support was assigned. As nucleotide sequecnes were not available for AfA, we used subtype information as provided by the cohort based on reverse transcriptase (RT) and protease. For Aquitaine, information on subtype was used where available, and otherwise considered as unknown. The Aquitaine subtypes were characterised locally using Blast analysis on Smartgene HIV module on two genes at least. In the analysis, we grouped HIV subtypes other than the four most common subtypes (B, C, A, G) as other (subtypes F, AD, AE, D, 06_CPX, 18_CPX, unknown). The appendix (p 1) provides further details.

### Definitions

Individuals who were prescribed raltegravir or elvitegravir before starting the DTG-based regimen were considered exposed to first-generation INSTIs. Viral load testing frequency was calculated for individuals with more than one year of follow-up before the Genotypic Resistance Test (GRT). We quantified HIV viral load as the area under the curve (AUC) of the log_10_-transformed viral load measurements from DTG initiation to the GRT sample date. To account for differences in detection limits, we set any viral load measurement below 50 to 0 copies/ml. For individuals who initiated ART with the DTG-based regimen, we excluded high viral loads at ART initiation by setting measurements within the first 180 days from the first HIV RNA measurement to 0. Time on DTG-based ART was calculated in years from DTG initiation to GRT. The ART regimen at GRT was considered the regimen an individual was taking 14 days before the test. If available, GRT results from earlier time points were used to assess prior NRTI resistance.

### Statistical analysis

We used descriptive statistics to present the characteristics of the study population and the different INSTI drug-resistance mutations. A negative binomial generalised linear model, adjusting for HIV subtype, exposure to first-generation INSTIs, and sex, was used to analyse the number of major and accessory INSTI drug-resistance mutations. We used ordinal logistic regression to identify risk factors for developing resistance, including cohort as a random effect. We considered variables based on availability and clinical relevance. We included sex, age at initiation and time on the DTG-based regimen, HIV subtype, type of ART (combination ART based on three drugs or more, DTG lamivudine dual therapy, other lamivudine dualtherapy, or monotherapy), exposure to first-generation INSTIs, HIV viral load, viral load testing frequency, and resistance to NRTIs. The missing data was included as a separate category if the sequencing did not cover the RT. All analyses were performed in R, version 4·0·5.

### Sensitivity analyses

We performed several sensitivity analyses. First, we replaced the NRTI resistance variable with the presence or absence of the M184V/I mutation (sensitivity analysis S1). Further, we performed logistic regression using the same covariables as in the main risk factor analysis, using susceptible versus any DTG resistance as the outcome (S2). We repeated the risk factor analyses excluding study participants where RT was not sequenced (S3). Given the widespread use of tenofovir-lamivudine-dolutegravir (TLD), we restricted the analysis of NRTIs to tenofovir and lamivudine and used the higher resistance level of the two to quantify NRTI resistance (S4). In the subset of people on a DTG + 2 NRTI regimen, we calculated NRTI resistance specific to the two NRTIs used in each participant (S5). The main analysis could not assess whether NRTI and NNRTI resistance mutations pre-existed or were acquired on DTG. Sensitivity analysis S6 restricted the study population to participants with available GRTs before experiencing virological failure on the DTG-containing regimen.

## Results

A total of 750 people met the eligibility criteria and were included in the analysis of mutations conferring resistance to DTG; 677 (90·3%) had more than one year of follow-up since starting the DTG-based regimen and were included in the analysis of risk factors for DTG resistance.

### Characteristics of the study population

The study participants included in the two analyses were similar (Table 1): most participants were men living with HIV subtype B who were on combination ART with three or more antiretroviral drugs. The median year of starting DTG was 2016. People were on DTG for a median of 16 and 1·7 years, and the median AUC of log_10_ viral load (copies/mL) accumulated during this period was around 3. The first GRT was performed in May 2013, and the last in January 2022. About a third of participants had previously been exposed to first-generation INSTIs. A total of 193 people did not have a CD4 measurement within a year of the GRT, 25 did not have any recorded HIV RNA measurements before the GRT, and in 74 people sequencing did not cover RT. The appendix (p 1) provides further details on the ART regimens.

**Table 1:**
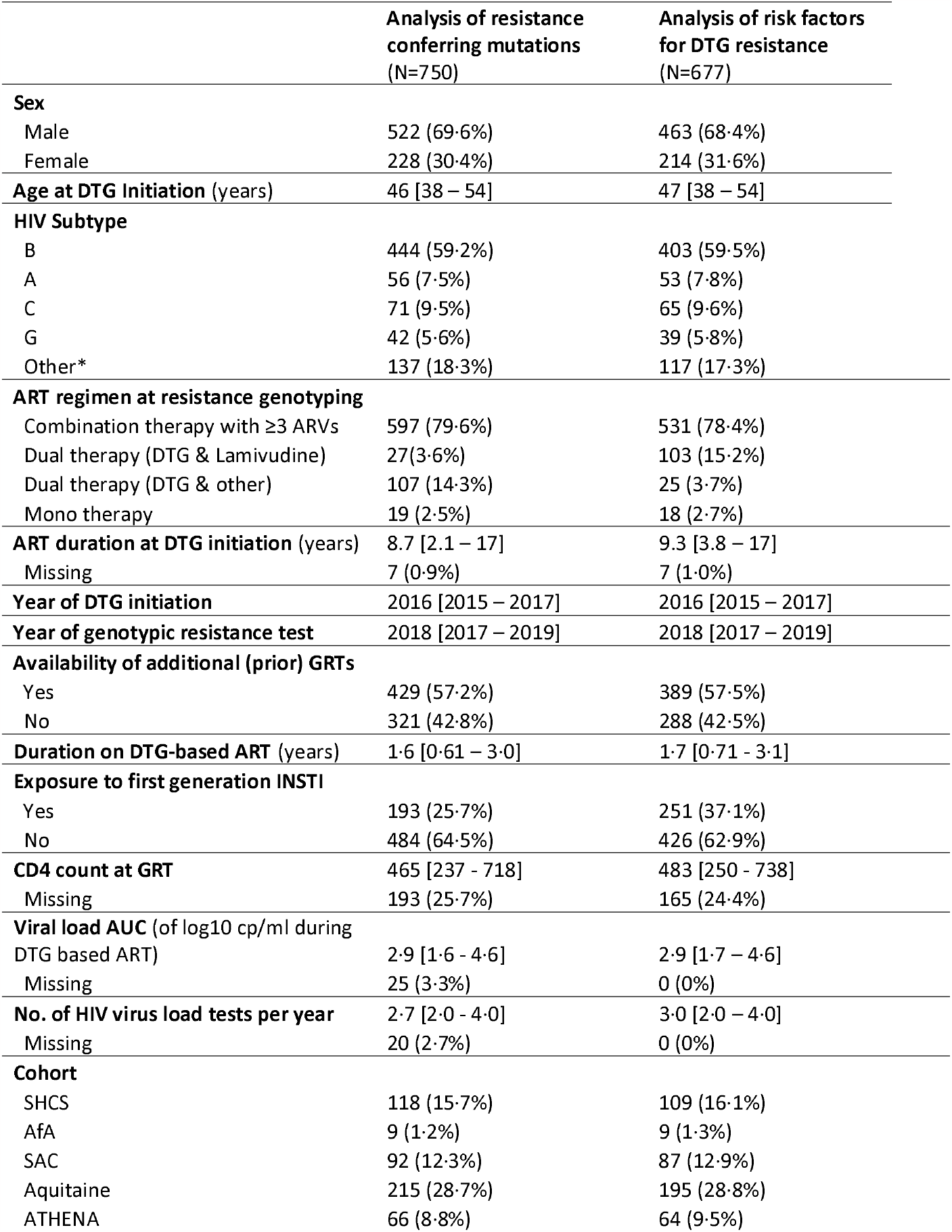
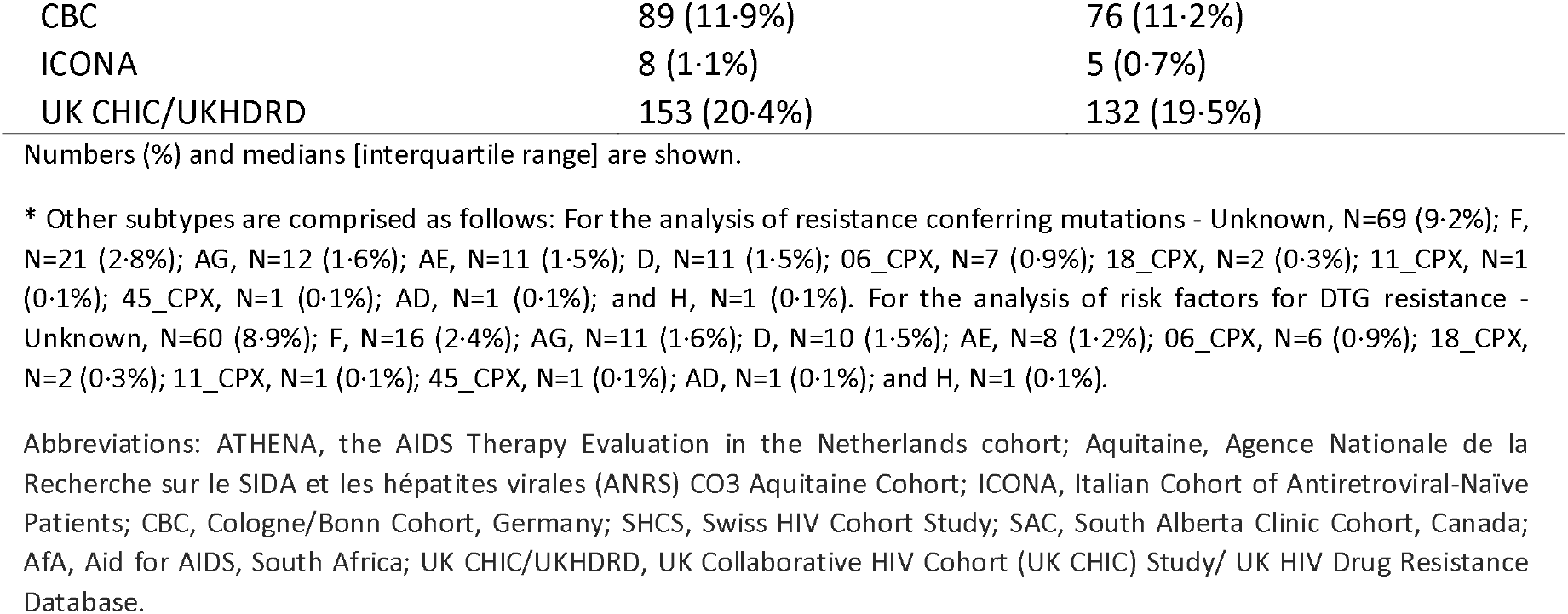
Demographics and clinical characteristics in the study population. People with virological failure on DTG-based ART with available genotypic resistance tests from eight observational HIV cohorts were included in the study. Study participants where clinical data was available for at least one year were considered for analysing risk factors for DTG resistance.

|  | Analysis of resistance<br>conferring mutations<br>(N=750) | Analysis of risk factors<br>for DTG resistance<br>(N=677) |
| --- | --- | --- |
| **Sex** |  |  |
| Male | 522 (69.6%) | 463 (68.4%) |
| Female | 228 (30.4%) | 214 (31.6%) |
| **Age at DTG Initiation (years)** | 46 [38 – 54] | 47 [38 – 54] |
| **HIV Subtype** |  |  |
| B | 444 (59.2%) | 403 (59.5%) |
| A | 56 (7.5%) | 53 (7.8%) |
| C | 71 (9.5%) | 65 (9.6%) |
| G | 42 (5.6%) | 39 (5.8%) |
| Other* | 137 (18.3%) | 117 (17.3%) |
| **ART regimen at resistance genotyping** |  |  |
| Combination therapy with ≥3 ARVs | 597 (79.6%) | 531 (78.4%) |
| Dual therapy (DTG & Lamivudine) | 27(3.6%) | 103 (15.2%) |
| Dual therapy (DTG & other) | 107 (14.3%) | 25 (3.7%) |
| Mono therapy | 19 (2.5%) | 18 (2.7%) |
| **ART duration at DTG initiation (years)** | 8.7 [2.1 – 17] | 9.3 [3.8 – 17] |
| Missing | 7 (0.9%) | 7 (1.0%) |
| **Year of DTG initiation** | 2016 [2015 – 2017] | 2016 [2015 – 2017] |
| **Year of genotypic resistance test** | 2018 [2017 – 2019] | 2018 [2017 – 2019] |
| **Availability of additional (prior) GRTs** |  |  |
| Yes | 429 (57.2%) | 389 (57.5%) |
| No | 321 (42.8%) | 288 (42.5%) |
| **Duration on DTG-based ART (years)** | 1.6 [0.61 – 3.0] | 1.7 [0.71 - 3.1] |
| **Exposure to first generation INSTI** |  |  |
| Yes | 193 (25.7%) | 251 (37.1%) |
| No | 484 (64.5%) | 426 (62.9%) |
| **CD4 count at GRT** | 465 [237 - 718] | 483 [250 - 738] |
| Missing | 193 (25.7%) | 165 (24.4%) |
| **Viral load AUC (of log10 cp/ml during DTG based ART)** | 2.9 [1.6 - 4.6] | 2.9 [1.7 – 4.6] |
| Missing | 25 (3.3%) | 0 (0%) |
| **No. of HIV virus load tests per year** | 2.7 [2.0 - 4.0] | 3.0 [2.0 – 4.0] |
| Missing | 20 (2.7%) | 0 (0%) |
| **Cohort** |  |  |
| SHCS | 118 (15.7%) | 109 (16.1%) |
| AfA | 9 (1.2%) | 9 (1.3%) |
| SAC | 92 (12.3%) | 87 (12.9%) |
| Aquitaine | 215 (28.7%) | 195 (28.8%) |
| ATHENA | 66 (8.8%) | 64 (9.5%) |
| CBC | 89 (11.9%) | 76 (11.2%) |
| ICONA | 8 (1.1%) | 5 (0.7%) |
| UK CHIC/UKHDRD | 153 (20.4%) | 132 (19.5%) |

Numbers (%) and medians [interquartile range] are shown.
\* Other subtypes are comprised as follows: For the analysis of resistance conferring mutations - Unknown, N=69 (9.2%); F, N=21 (2.8%); AG, N=12 (1.6%); AE, N=11 (1.5%); D, N=11 (1.5%); 06\_CPX, N=7 (0.9%); 18\_CPX, N=2 (0.3%); 11\_CPX, N=1 (0.1%); 45\_CPX, N=1 (0.1%); AD, N=1 (0.1%); and H, N=1 (0.1%). For the analysis of risk factors for DTG resistance - Unknown, N=60 (8.9%); F, N=16 (2.4%); AG, N=11 (1.6%); D, N=10 (1.5%); AE, N=8 (1.2%); 06\_CPX, N=6 (0.9%); 18\_CPX, N=2 (0.3%); 11\_CPX, N=1 (0.1%); 45\_CPX, N=1 (0.1%); AD, N=1 (0.1%); and H, N=1 (0.1%).
Abbreviations: ATHENA, the AIDS Therapy Evaluation in the Netherlands cohort; Aquitaine, Agence Nationale de la Recherche sur le SIDA et les hépatites virales (ANRS) CO3 Aquitaine Cohort; ICONA, Italian Cohort of Antiretroviral-Naïve Patients; CBC, Cologne/Bonn Cohort, Germany; SHCS, Swiss HIV Cohort Study; SAC, South Alberta Clinic Cohort, Canada; AfA, Aid for AIDS, South Africa; UK CHIC/UKHDRD, UK Collaborative HIV Cohort (UK CHIC) Study/ UK HIV Drug Resistance Database.

### INSTI mutations and DTG resistance

At least one major or accessory INSTI DRM was found in 100 (13·3%) of the 750 study participants; 21 (2·8%) had more than one mutation. Most (713; 95·1%) study participants were fully susceptible to DTG, with potential low, low, intermediate, and high levels of DTG resistance being observed in 8 (1·1%), 5 (0·7%), 18 (2·4%) and 6 (0·8%), respectively (Figure 1). The INSTI resistance mutations observed in all enrolled people with a DTG resistance score > 0 are shown in the appendix (p 2).

**Figure 1:**
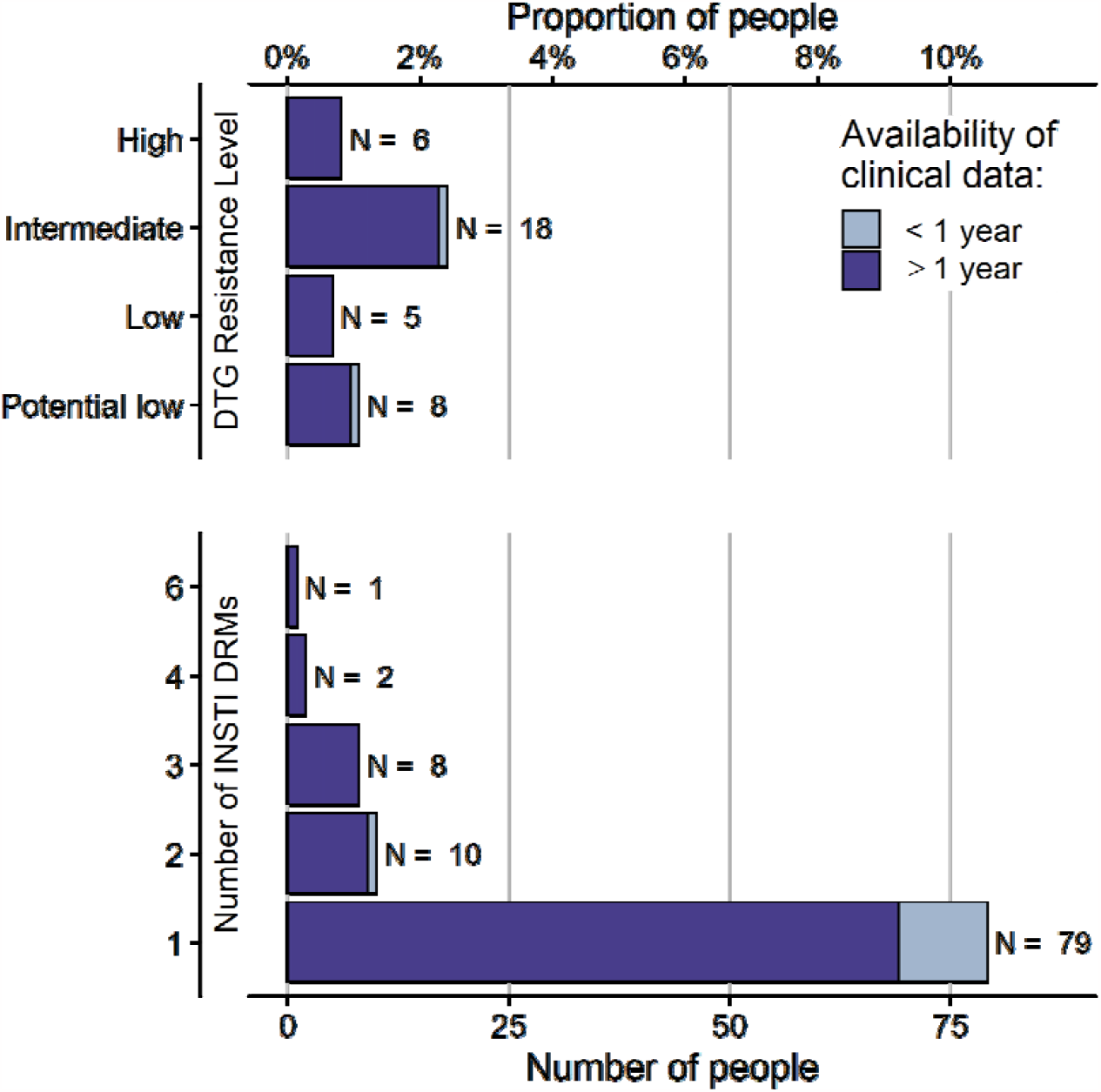
Prevalence of DTG resistance and INSTI DRMs. Genotypic resistance tests of 750 people with genotypic resistance testing on DTG-based ART were analysed using the Stanford resistance algorithm to determine INSTI DRMs and resistance level to DTG. Both major and accessory INSTI DRMs were considered for the number of INSTI DRMs. People with no INSTI DRMs (N = 650, 86·7%), and who are susceptible to DTG (N = 713, 95·1%) are not displayed.

The most common major INSTI DRM was R263K (N=11), which only once occurred with another major INSTI DRM (appendix p 2). Other common major mutations included the G140 (N = 10), N155 (N = 9), Q148 (N = 8), and the E138 mutation (N = 7). The G118R mutation, which has the strongest impact on susceptibility to DTG, was only observed three times. Among accessory DRMs, E157 (N = 25) and T97 (N = 20) were the most common. The distribution of INSTI resistance mutations was similar in people previously exposed to first-generation INSTIs and those not exposed (Figure 2). There was no statistically significant association of specific DRMs with first-generation INSTI experience. For HIV subtype, we found a significant association for the accessory INSTI DRM T97 (see appendix p 3). This DRM occurred in 6 of 56 (10·7%) people with HIV subtype A, 4 of 42 (9·5%) people with subtype G, 7 of 444 (1·6%) people with subtype B, and 0 of 71 people with HIV subtype C.

**Figure 2:**
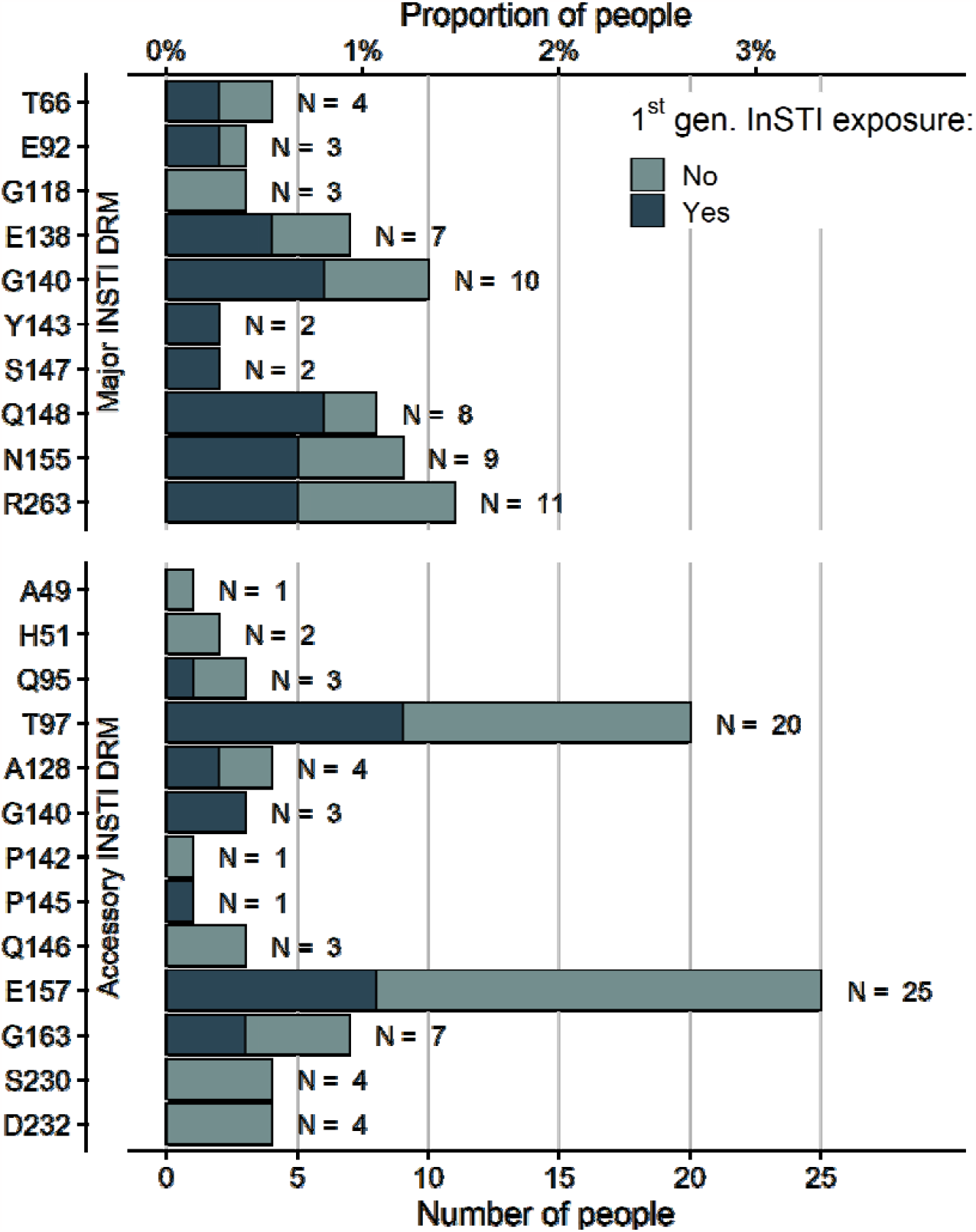
INSTI drug resistance mutations found in 750 people experiencing virologic failure on a DTG-based regimen. Drug resistance mutations were classified as major and accessory according to the Stanford resistance database^27^. Bars are coloured by previous history of first generation INSTIs (raltegravir, elvitegravir).

The results from the negative binomial model of the number of mutations showed little evidence of a difference between HIV subtypes. The number of INSTI DRMs was higher in first-generation INSTI-exposed people (adjusted RR 1·59, 95% CI 1·03 to 2·48) (Figure 3). This association became even stronger when considering the number of major INSTI DRMs (adjusted RR 2·60, 95% CI 1·30 to 5·31) (see appendix p 4 for further details).

**Figure 3:**
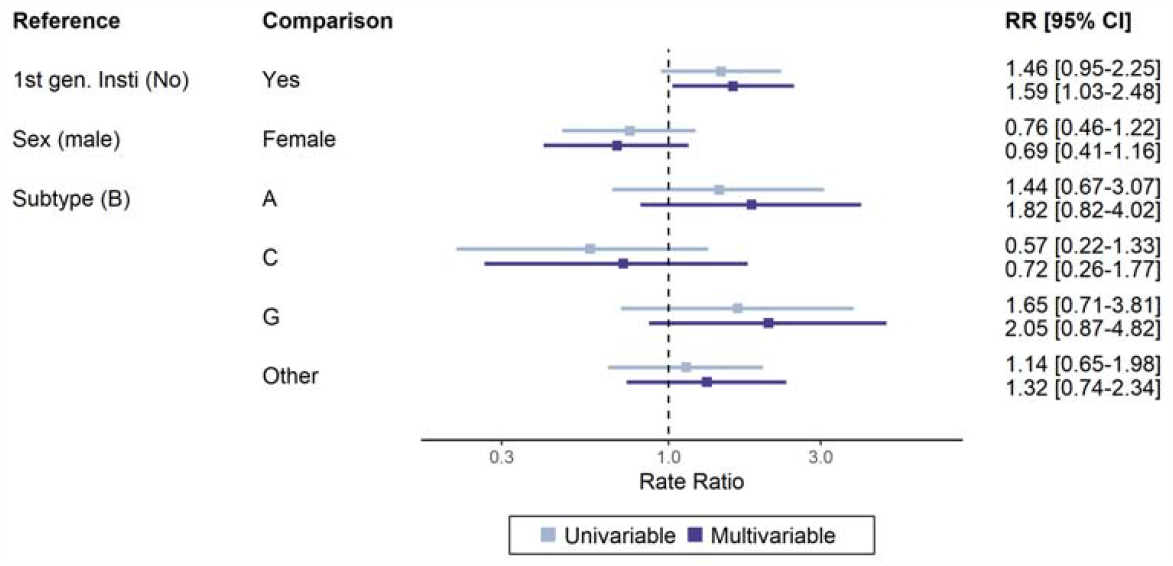
Rate ratio for the number of INSTI DRMs. A negative binomial generalised linear model was fit to the number of major and accessory INSTI DRMs in 750 people with virological failure on DTG-based ART. The plot shows uni- and multivariable point estimates and 95% confidence intervals of rate ratios.

The prevalence of resistance mutations (low, intermediate or high) to NRTIs and NNRTIs was substantially higher in the presence of DTG resistance (Table 2). Among GRTs with coverage of the RT, the prevalence of at least low level NRTI resistance was 10·1% overall (66 of 654) but 31·8% (7 of 22) among those with DTG resistance. The corresponding figures for NNRTI resistance were 12·8% (84 of 654) and 50% (11 of 22).

**Table 2:** Resistance levels to DTG, non-nucleoside reverse transcriptase inhibitors and nucleotide reverse transcriptase inhibitors. The number and percentage of people with corresponding drug resistance levels are given for the entire study population. NRTI resistance level is based on median resistance score to ABC, AZT, XTC and TDF/TAF. NNRTI resistance level is based on median resistance score to EFV, ETR, NVP, and RPV.

|  | **DTG resistance level** |  |
| --- | --- | --- |
|  | **Susceptible & Potential Low**<br>**(N=721)** | **Low, Intermediate, High**<br>**(N=29)** |
| **NRTI resistance level** |  |  |
| Susceptible | 574 (79.6%) | 13 (44.8%) |
| Potential Low | 14 (1.9%) | 2 (6.9%) |
| Low | 14 (1.9%) | 1 (3.4%) |
| Intermediate | 13 (1.8%) | 2 (6.9%) |
| High | 39 (5.4%) | 4 (13.8%) |
| RT not covered in GRT | 67 (9.3%) | 7 (24.1%) |
| **NNRTI resistance level** |  |  |
| Susceptible | 543 (75.3%) | 11 (37.9%) |
| Potential Low | 27 (3.7%) | 0 (0%) |
| Low | 21 (2.9%) | 0 (0%) |
| Intermediate | 39 (5.4%) | 4 (13.8%) |
| High | 24 (3.3%) | 7 (24.1%) |
| RT not covered in GRT | 67 (9.3%) | 7 (24.1%) |

### Risk factors for DTG resistance

The risk of DTG resistance was higher on DTG monotherapy compared to combination ART with >3 drugs (adjusted odds ratio [aOR] 37·25, 95% CI 11·17 to 124·2) and for DTG lamivudine dual regimen (aOR 6·59, 95% CI 1·70 to 25·55) (Figure 4). The risk of resistance was also increased in the presence of a potential low/low level of NRTI resistance (aOR 4·62, 95% CI 1·24 to 17·2) or intermediate/high level (aOR 7·01, 95% CI 2·52 to 19·48), compared to no NRTI resistance. Non-B HIV subtypes tended towards higher resistance levels, mainly driven by subtype A (aOR 3·27, 95% CI 0·90 to 11·87) (appendix p 5). There was a trend for an association of viral load with DTG resistance (aOR 1·42, 95% CI 0·92 to 2·19 per standard deviation of the log_10_ virus load area under the curve).

**Figure 4:**
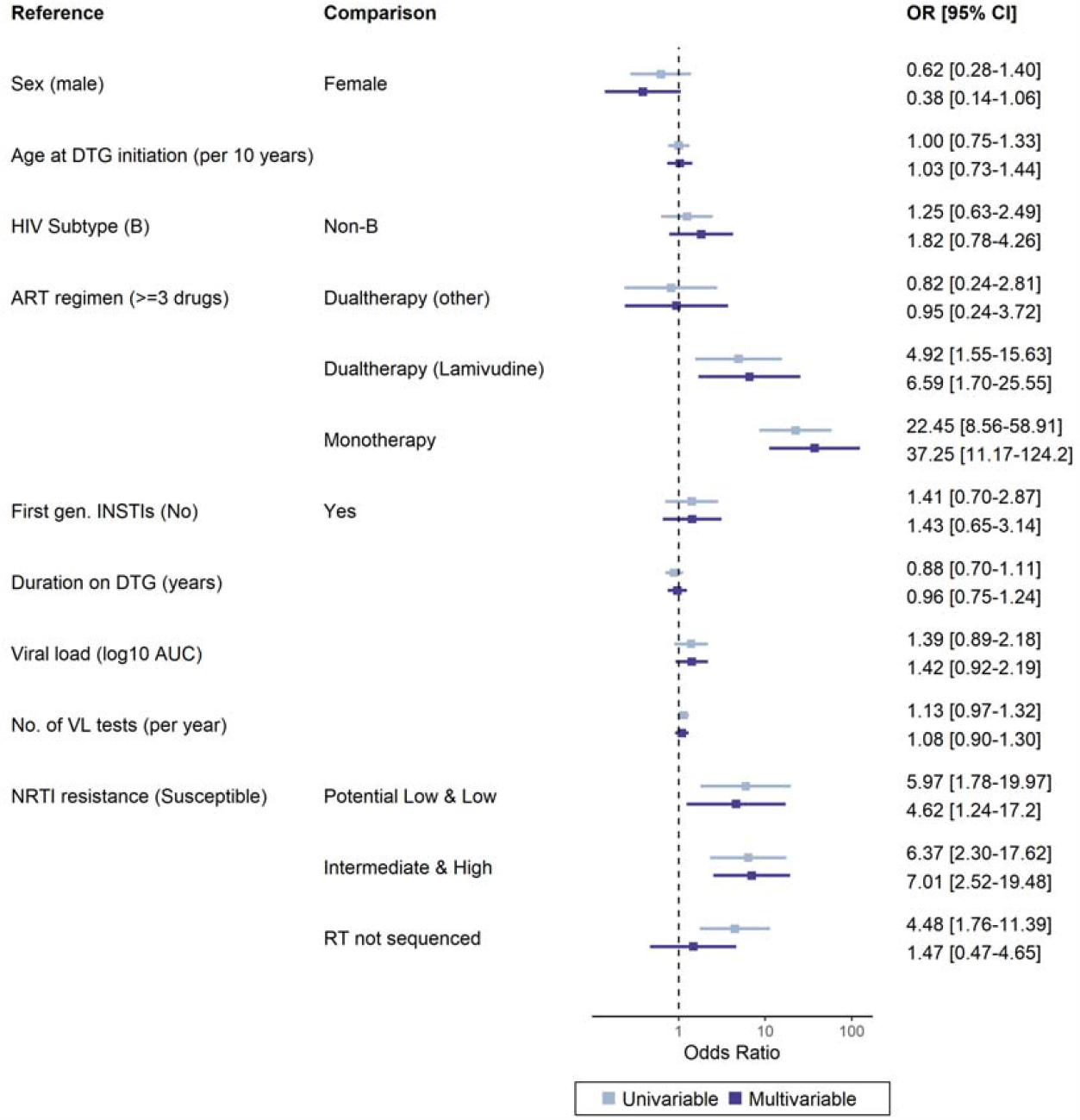
Odds ratios for DTG resistance levels with 95% confidence intervals from uni- and multivariable ordinal logistic models for genotypic DTG resistance. Cohorts were included as random effect. DTG resistance levels in people with virological failure on DTG-based ART were assessed using the Stanford resistance algorithm.

### Sensitivity analyses

The results of the risk factor analyses were similar when replacing the NRTI resistance variable with the M184V/I mutation (sensitivity analysis S1, appendix p 7) or when analysing susceptible versus any DTG resistance as the outcome in a logistic regression (S2, appendix p 8). The exclusion of 64 individuals with missing RT sequences allowed the inclusion of both NRTI and NNRTI resistance in the model. The results for NRTI resistance were similar, and intermediate/high-level NNRTI resistance was also associated with DTG resistance (adjusted OR 4·07, 95% CI 1·07 – 15·5) (S3, appendix p 9). The analysis restricted to lamivudine and tenofovir (S4, appendix p 10) confirmed that DTG resistance was associated with potential low/low and intermediate/high-level resistance to these NRTI drugs. Similarly, when restricting the analysis to people on a DTG regimen with two NRTI backbones, we found similar results for the specific NRTIs (S5, appendix p 11). Finally, in the last sensitivity analysis (S6) we used data on pre-existing NRTI resistance and found that DTG resistance was associated with prior intermediate/high NRTI resistance (appendix p 12).

## Discussion

In this collaborative analysis of eight large cohort studies, we identified INSTI DRMs in 100 of 750 (13·3%) PLHIV who experienced virologic failure on DTG-based ART, and resistance to DTG according to the Stanford algorithm was present in 37 (4·9%) individuals. DTG resistance was associated with DTG monotherapy, lamivudine DTG dual therapy, resistance to RT inhibitors, non-B subtype, and HIV viral replication but not with previous first-generation INSTI exposure. A wide range of INSTI DRMs was present. The polymorphic accessory INSTI DRM T97A was detected more frequently in subtypes A and G (compared to subtypes B and C), consistent with previously reported data.^30^ The major INSTI DRMs at positions 140 and 148 were detected in 5 out of 6 people with high-level DTG resistance, all of whom were first-generation INSTI experienced.

DTG monotherapy, DTG lamivudine dual therapy and resistance to the NRTI backbone were most strongly associated with DTG resistance in our study. The complete sequence analysis, which allowed us to distinguish between NRTI and NNRTI resistance, suggests that the association may be mediated via NRTI resistance. It was robust when considering only 3TC and TDF resistance in a sensitivity analysis. As the main analysis was cross-sectional, it did not allow the determination of the timing of NRTI resistance relative to DTG resistance. However, an additional analysis in people with prior resistance tests suggests that NRTI resistance may often have predated DTG resistance. These results indicate that resistance to NRTI backbone drugs from previous regimens may have promoted the emergence of DTG resistance. However, its also possible that prior NRTI resistance reflects adherence issues, which may facilitate the emergence of DTG resistance.

The strong associations of DTG resistance with DTG monotherapy and NRTI resistance are consistent with previous findings,^10,31,32^ but appear to contradict the results from the NADIA trial, which found no evidence that the efficacy of DTG-based ART is affected by resistance to the NRTI backbone.^10^ However, it should be noted that the NADIA trial examined the risk of virological failure in 235 PLHIV randomised to DTG, resulting in a small number of treatment failures (n=24), while our study focused on the risk of resistance among people who experienced virological failure. Thus, for DTG-based regimens, NRTI resistance may not substantially affect the risk of treatment failure but still increase the risk of resistance in case of failure. Research on other drug classes^33^ has shown that drug regimens with high and low genetic barriers can have similar failure rates but different probabilities of acquiring resistance.

Our study contributes new information on DTG resistance in PLHIV receiving different DTG-based ART regimens by examining risk factors for DTG resistance in real-world cohort data from different settings. The cohort collaboration resulted in a large dataset of GRT results in people who experienced virological failure on DTG. Our results are central to informing HIV treatment and monitoring policies in the context of the continued expansion of DTG-based treatment regimens. The pooling of data from diverse routine clinical cohorts also has limitations. The data come from individuals receiving any DTG-based regimen in a wide range of different clinical care settings. We only included people who experienced virological failure and had a GRT, but policies and practices regarding when and for whom GRT is done likely differed between cohorts. In our regression models, we accounted for this heterogeneity between cohorts by including cohort as a random effect, but confounding by cohort may still have affected our results. Further, the personalised approach to ART and HIV care in the European settings will not be generalisable to other settings, particularly low- and middle-income countries.

A further limitation of our study is the dominance of HIV subtype B, which was expected considering that our study population is comprised mainly of PLHIV from European countries, where subtype B predominates. More data from people with non-B subtypes are needed, and such a study is ongoing within the framework of the International epidemiology Databases to Evaluate AIDS (IeDEA).^26^ In this study, individuals experiencing failure on DTG-based ART are prospectively enrolled in around forty sites across sub-Saharan Africa, South America, and Asia. Furthermore, the WHO plans to launch sentinel surveys of acquired HIV resistance to DTG among people receiving DTG-based ART.^34^ We could not assess adherence or drug interactions with rifampicin, which may influence the emergence of DTG resistance.^35^ Adherence and rifampicin use were not recorded consistently and comparably in the participating cohorts. In our study population, the DTG-based regimens were too heterogenous to allow investigating DTG resistance outcomes of specific regimens and treatment histories. Lastly, there is growing evidence that mutations outside integrase may confer DTG resistance^36–38^. Our study was based on pol sequences, which did not allow us to investigate these mutations.

The associations we found with DTG resistance, resistance to NRTI backbone drugs, HIV-1 subtype, and unsuppressed virus load have important implications for ensuring the long-term sustainability of ART. While INSTI resistance was rare in our population; it is still a concern. Firstly, the duration of DTG therapy and the duration of viraemia whilst receiving DTG was relatively short in our population: the median time on DTG was less than 2 years and drug resistance might emerge more frequently in settings where individuals remain viraemic for a longer time on DTG regimens. This could happen in resource-limited settings where guidelines recommend not switching from DTG-based therapy unless multiple viral loads >1000 copies/mL have been documented and where delays in regimen switching are common.^39^ Secondly, the strong association of DTG resistance with NRTI resistance suggests that the risk of resistance might be higher in people with previous failure on NNRTI-based first-line therapy, among whom the prevalence of NRTI resistance is much higher than in our study population. The WHO guidelines recommend DTG in 1st-, 2nd- and 3rd-line ART. This multiplicity of roles combined with the recycling of drugs and limited access to viral load and drug resistance testing will facilitate the emergence of DTG resistance. Finally, even a relatively low level of acquired DTG resistance in the millions of people receiving DTG-based ART could lead to rising levels of transmitted INSTI resistance, which could affect both treatment and prevention

In conclusion, our study underlines the importance of routine viral load monitoring and resistance testing, especially in treatment-experienced people, to prevent resistance both at the individual patient and the population level and thereby ensure the long-term sustainability of ART.

## Data Availability

Data underlying the figures and tables reported in this article may be shared following publication of this article. Researchers may submit a methodologically sound proposal for the use of these data to the corresponding author. De-identified data of individual study participants cannot be made available as they are subject to the respective observational HIV cohorts.

## Authors’ contributions

Conceptualisation (HFG, JACS, RL, ME, RK), Data curation (TL, SH, SI, HO), Methodology (TL, CS, RK ME, JACS), Formal analysis & Validation (TL, RK), Investigation (TL, SH, SI), Project administration (SH, SI), Resources (HO, KK, JM, AvS, MS, AAM, JG, CS, GM), Software (TL, KK), Supervision (HFG, JACS, RL, ME, RK), Visualisation (TL), Writing – original draft (TL, RL, ME, RK), Writing – review & editing (All authors).

TL and RK have directly accessed and verified the underlying data reported in the manuscript. RL, ME and RK contributed equally.

## Acknowledgements

We would like to acknowledge Anthony Hauser, Suraj Balakrishna, and Marius Zeeb for helpful discussions on data analysis. This study was supported by the National Institute Of Allergy And Infectious Diseases of the National Institutes of Health under Award Number R01AI152772 and the Swiss National Science Foundation (32FP30_207285, 324730_207957). The participating cohorts or cohort collaborations were funded by the Swiss National Science Foundation (33CS30_201369) and the Yvonne Jacob Foundation (for the SHCS), the UK Medical Research Council (grant numbers G0000199, G0600337, G0900274, and M004236/1; for the UK Collaborative HIV Cohort), the National Agency for AIDS Research (France REcherche Nord&Sud Sida-hiv Hépatites), the French Agency for Research on AIDS and Viral Hepatitis ⍰Emerging Infectious Diseases (ANRS⍰MIE) and the CHU de Bordeaux (for the ANRS CO3 Aquitaine-AquiVIH-NA cohort), the Dutch Ministry of Health (for the ATHENA cohort), the German Center for Infection Research (8018704707) (for the CBC), ICONA Foundation is supported by unrestricted grants from BMS, Gilead Sciences, Janssen, MSD and ViiV Healthcare. AFA is supported via IeDEA-SA by the U.S. National Institutes of Health’s National Institute of Allergy and Infectious Diseases, the Eunice Kennedy Shriver National Institute of Child Health and Human Development, Division of Cancer Epidemiology and Genetics, National Cancer Institute, the National Institute of Mental Health, the National Institute on Drug Abuse, the National Heart, Lung, and Blood Institute, the National Institute on Alcohol Abuse and Alcoholism, the National Institute of Diabetes and Digestive and Kidney Diseases and the Fogarty International Center under Award Number U01AI069924. The ART-CC is funded by the US National Institute on Alcohol Abuse and Alcoholism (U01-AA026209). The content is solely the responsibility of the authors and does not necessarily represent the official views of the National Institutes of Health.

