## Supplemental appendix for "HIV-1 drug resistance in people on dolutegravir-based ART: Collaborative analysis of cohort studies"

**On HIV subtyping**

For AfA and Aquitaine, subtyping was performed by them. In AfA, all subtype were C, except one denoted as ?, which we assign to Unknown. For Aquitaine we assigned subtypes as follows: A: A1; B: B; C: C; AE: CRF01_AE; AG: CRF02_AG; 06_CPX: CRF06_cpx; 11_CPX: CRF11_cpx; 45_CPX: CRF45_cpx; D: D; F: F1, F2; G: G; Unknown: Non B non précisé, UNKNOWN.

For all other cohorts, HIV subtypes were determined on the integrase sequence using COMET (COntext-based Modeling for Expeditious Typing)^28^ and REGA.^29^ HIV subtypes where neither algorithm had support >= 50 were considered to be Unknown. Subtypes with support >= 50 were combined as follows for the two algorithms:

Comet subtypes with support >= 50: A: A1 and A6; AE: 01_AE; B: B; C: C; D: D; F: F1; G: G; H: H.

Rega subtypes with support >= 50: 18_CPX: HIV-1 CRF 18_cpx; A: HIV-1 Subtype A (A1), HIV-1 Subtype A (A1)-like, and HIV-1 Subtype A (A1), potential recombinant; AE: HIV-1 Subtype A (01_AE); B: HIV-1 Subtype B; C: HIV-1 Subtype C; D: HIV-1 Subtype D; F: HIV-1 Subtype F (F1); G: HIV-1 Subtype G and HIV-1 Subtype G, potential recombinant; H: HIV-1 Subtype H.

In case of differing results, the subtype with the higher support was taken.

**ART Regimen**

ART regimen 14 days before genotypic resistance testing were considered. In case of genotypic resistance testing within two months after DTG-based ART ended, we considered the ART regimen 14 days before the end of the DTG-based regimen. ART regimen for UK Chic were provided by the cohort.

Supplemental table 1: **ART regimens at genotypic resistance testing**. Regimens 14 days before the sampling date for resistance testing are considered. We grouped regimen as follows: Regimen with three ARVs or more not containing PI are considered as DTG + ≥2 NRTI. Next, DTG regimen containing PIs are considered as DTG + PI ± Other. Next, DTG regimen containing NNRTI are considered as DTG + NNRTI ± Other. Next, DTG regimen with two ARVs, the second being an NRTI, are considered as DTG + NRTI. Next, DTG regimen containing no other ARVs are considered DTG monotherapy. DTG regimen not described by the terms above are considered as Other.

| **Regimen** | **Number of study participants N, (%)** | **Most common ARV combinations**  **(N > 10)** |
| --- | --- | --- |
| DTG + ≥2 NRTI | 475 (63·3%) | 3TC, ABC, DTG (N = 295), DTG, FTC, TDF (N = 114), DTG, FTC, TAF (N = 43) |
| DTG + PI ± Other | 154 (20·5%) | DRV, DTG, RTV (N = 36), DRV, DTG, FTC, RTV, TDF (N = 11) |
| DTG + NNRTI ± Other | 66 (8·8%) | DTG, RPV (N = 42) |
| DTG + NRTI | 33 (4·4%) | 3TC, DTG (N = 27) |
| DTG monotherapy | 19 (2·5%) | DTG (N = 19) |
| Other | 3 (0·4%) |  |

**List of INSTI DRMs observed in all enrolled people**

Supplemental table 2: **INSTI DRMs in people genotypic resistance testing on DTG-based ART by DTG resistance level**. DRMs and resistance to DTG were determined using the Stanford resistance algorithm V9.0. In the DTG RESIST study population, 650/750 people did not have INSTI DRMs.

| **DTG resistance level** | **No. of  people (%)** | **Major INSTI DRMs** | **Accessory INSTI DRMs** | **DTG resistance  score** |
| --- | --- | --- | --- | --- |
| **High** |  |  |  |  |
|  | 1 (0·1%) | E138K,G140S,Q148H,N155H | - | 105 |
|  | 1 (0·1%) | G140S,Q148H,N155H | - | 75 |
|  | 1 (0·1%) | G118R,E138K | - | 70 |
|  | 3 (0·4%) | G140S,Q148H | T97A | 60 |
| **Intermediate** | |  |  |  |
|  | 2 (0·3%) | G118R | - | 50 |
|  | 1 (0·1%) | G140GS,Q148R | D232N | 45 |
|  | 1 (0·1%) | E138EK,S147SG,N155NH | Q95QK,T97TA,E157EQ | 40 |
|  | 1 (0·1%) | E138K,S147G,N155H | T97A | 40 |
|  | 1 (0·1%) | E92EG,R263RK | - | 30 |
|  | 1 (0·1%) | E92Q,N155H | - | 30 |
|  | 1 (0·1%) | N155NH | Q146QR,S230SR | 30 |
|  | 1 (0·1%) | R263K | A49G,D232DN | 30 |
|  | 6 (0·8%) | R263K | - | 30 |
|  | 3 (0·4%) | R263RK | - | 30 |
| **Low** |  |  |  |  |
|  | 1 (0·1%) | E138EK,G140GEKR | - | 20 |
|  | 1 (0·1%) | N155H | H51HY,T97A | 20 |
|  | 1 (0·1%) | - | E157EQ,S230R | 20 |
|  | 1 (0·1%) | - | S230R | 20 |
|  | 1 (0·1%) | - | S230R,D232N | 20 |
| **Potential low** | |  |  |  |
|  | 1 (0·1%) | E138EK | D232DN | 10 |
|  | 1 (0·1%) | E138EK | - | 10 |
|  | 3 (0·4%) | G140R | - | 10 |
|  | 1 (0·1%) | N155H | G163R | 10 |
|  | 1 (0·1%) | N155H | - | 10 |
|  | 1 (0·1%) | - | H51HY | 10 |
| **Susceptible** | |  |  |  |
|  | 2 (0·3%) | T66I | - | 5 |
|  | 1 (0·1%) | T66TI | - | 5 |
|  | 1 (0·1%) | Y143C | - | 5 |
|  | 1 (0·1%) | Y143YC | - | 5 |
|  | 1 (0·1%) | E92E_P | - | 0 |
|  | 2 (0·3%) | - | A128AT | 0 |
|  | 1 (0·1%) | - | A128T | 0 |
|  | 1 (0·1%) | - | A128T,G140GE | 0 |
|  | 3 (0·4%) | - | E157EQ | 0 |
|  | 20 (2·7%) | - | E157Q | 0 |
|  | 1 (0·1%) | - | G140G_S | 0 |
|  | 1 (0·1%) | - | G163GRS | 0 |
|  | 1 (0·1%) | - | G163K | 0 |
|  | 4 (0·5%) | - | G163R | 0 |
|  | 1 (0·1%) | - | P142T | 0 |
|  | 1 (0·1%) | - | P145P_H | 0 |
|  | 1 (0·1%) | - | Q146L | 0 |
|  | 1 (0·1%) | - | Q146R | 0 |
|  | 2 (0·3%) | - | Q95K | 0 |
|  | 12 (1·6%) | - | T97A | 0 |
|  | 2 (0·3%) | - | T97TA | 0 |
|  | 1 (0·1%) | Q148Q_E | G140G_R | 0 |
|  | 1 (0·1%) | Q148Q_R | - | 0 |
|  | 1 (0·1%) | T66A | - | 0 |

**Testing for association of INSTI DRM with HIV subtype and first-generation INSTI experience**

Supplemental table 3: **Association of INSTI DRMs with HIV subtype and first generation INSTI exposure**. We considered HIV subtypes B, C, A, G, and Other. Statistical tests were done using a fisher’s exact test. P-values were adjusted for multiple testing by Benjamini & Hochberg correction. The DRM at T97 was found to be associated with HIV subtype (bold): it occurred in 6 / 56 (10·7%) of people with HIV subtype A, 4 / 42 (9·5%) of people with subtype G, 7 / 444 (1·6%) of people with subtype B, and 0 / 71 people with HIV subtype C.

|  |  |  | **HIV subtype** | **First generation INSTI** |
| --- | --- | --- | --- | --- |
|  |  | **Observations** | Adjusted p-value | Adjusted p-value |
| **Major INSTI DRMs** | | |  |  |
|  | 66 | 4 | 1·000 | 0·790 |
|  | 92 | 3 | 1·000 | 0·682 |
|  | 118 | 3 | 1·000 | 0·790 |
|  | 138 | 7 | 1·000 | 0·682 |
|  | 140 | 10 | 0·511 | 0·682 |
|  | 143 | 2 | 1·000 | 0·682 |
|  | 147 | 2 | 1·000 | 0·682 |
|  | 148 | 8 | 1·000 | 0·509 |
|  | 155 | 9 | 1·000 | 0·682 |
|  | 263 | 11 | 0·367 | 0·790 |
| **Accessory INSTI DRMs** | | |  |  |
|  | 49 | 1 | 0·503 | 1·000 |
|  | 51 | 2 | 0·195 | 0·790 |
|  | 95 | 3 | 1·000 | 1·000 |
|  | 97 | 20 | **0·009** | 0·682 |
|  | 128 | 4 | 0·259 | 0·790 |
|  | 140 | 3 | 1·000 | 0·509 |
|  | 142 | 1 | 1·000 | 1·000 |
|  | 145 | 1 | 1·000 | 0·682 |
|  | 146 | 3 | 1·000 | 0·790 |
|  | 157 | 25 | 0·503 | 0·958 |
|  | 163 | 7 | 1·000 | 0·851 |
|  | 230 | 4 | 1·000 | 0·682 |
|  | 232 | 4 | 1·000 | 0·682 |

**Negative binomial model for number of major INSTI DRMs**

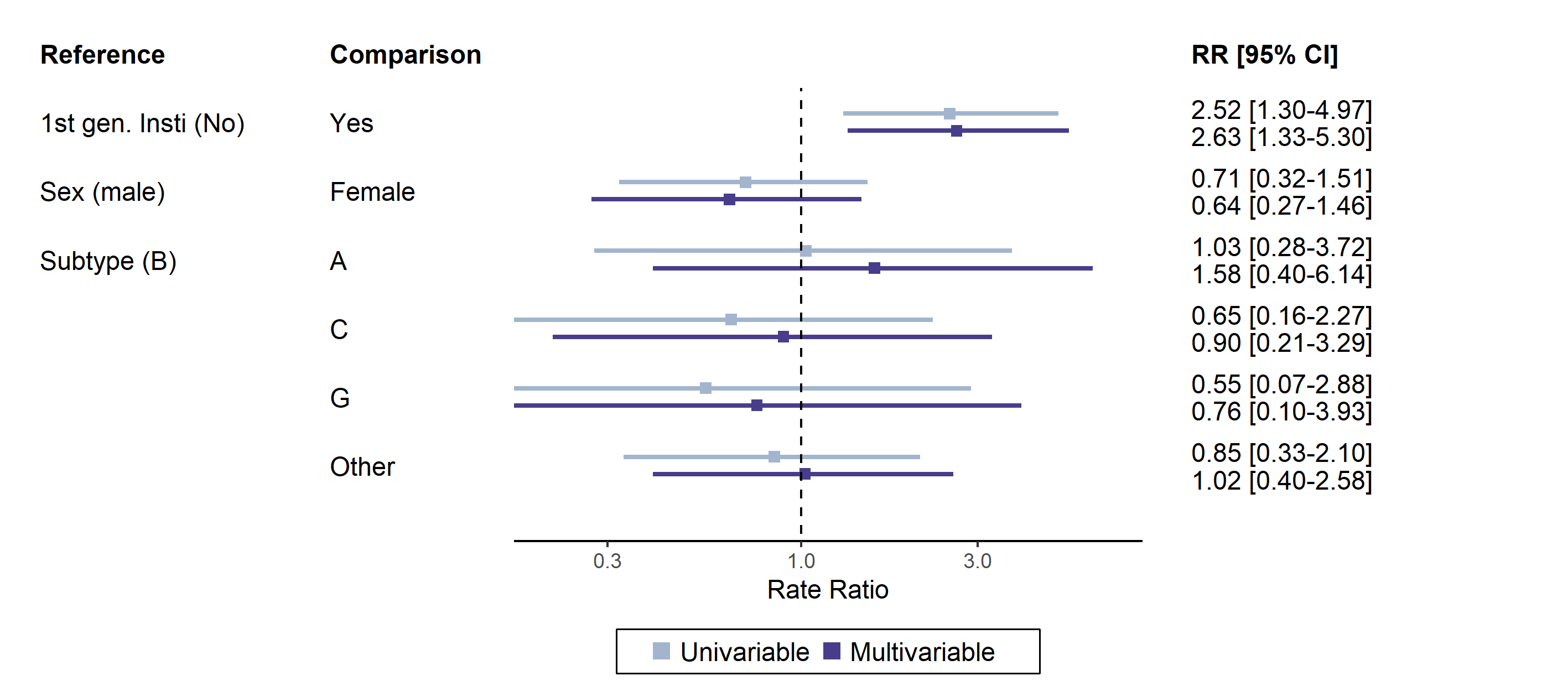

Supplemental figure 1: **Rate ratio for number of major INSTI DRMs**. A negative binomial generalised linear model was fit to the number of major INSTI DRMs in 750 people with virological failure on DTG-based ART. While 100 people had at least one INSTI DRM, only 42 had at least one major INSTI DRM. The figure shows uni- and multivariable point estimates and 95% confidence intervals of rate ratios.

**Risk factors for DTG resistance including HIV subtype B, A, C, G, and other**

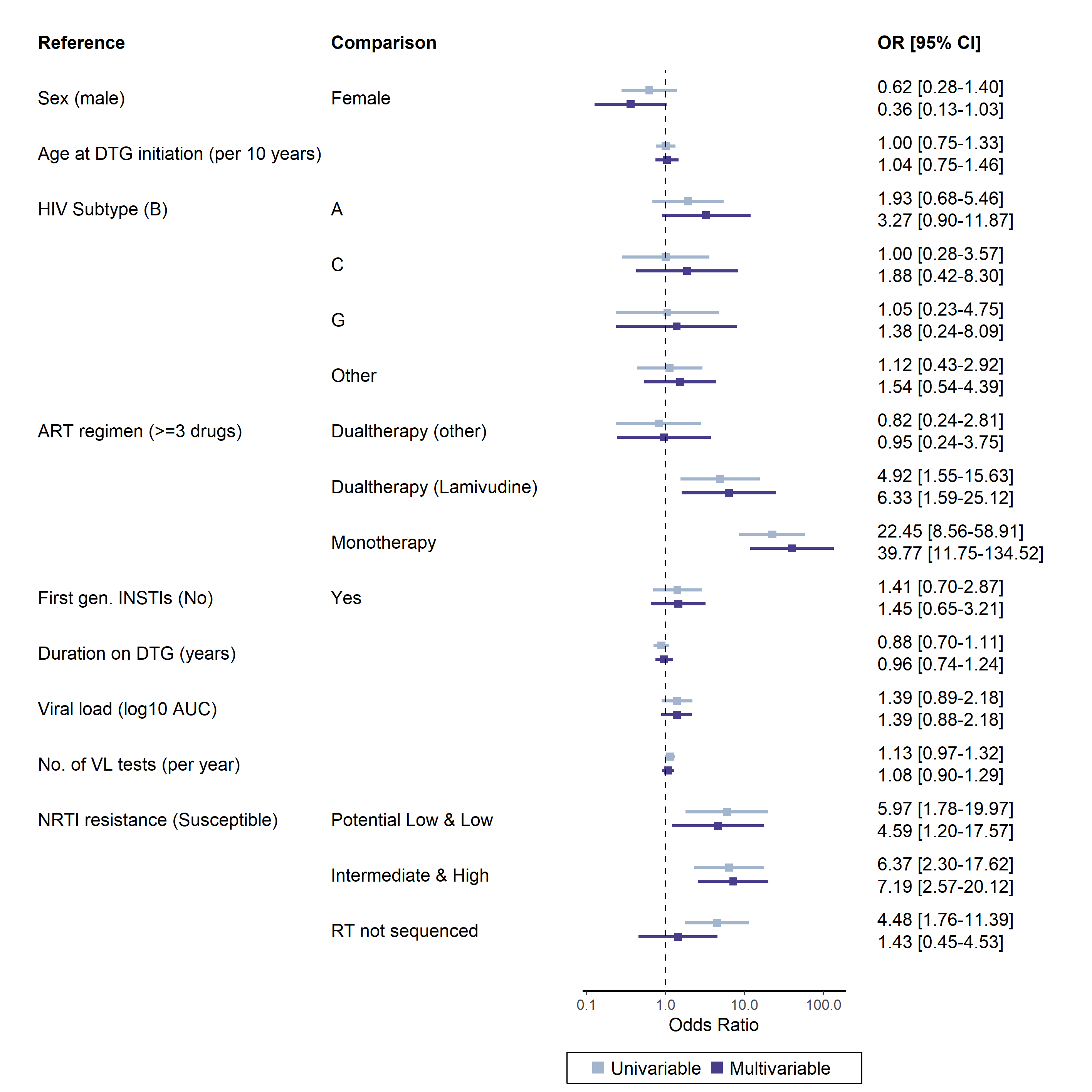

Supplemental figure 2: **Odds ratios for DTG resistance levels with 95% confidence intervals from uni- and multivariable ordinal logistic models for genotypic DTG resistance**. Cohorts were included as random effect. In this analysis, Non-B HIV subtypes were further separated.

**Sensitivity analyses**

We performed the following sensitivity analyses:

Sensitivity analysis S1: Using M184I/V as covariable in the risk factor analysis. As hallmark NRTI drug resistance mutation, in S1 we consider the M184I/V mutation as covariable in the risk factor analysis.

Sensitivity analysis S2: Using logistic regression for the risk factor analysis. In S2, we confirm the findings from the ordinal logistic regression using a simpler statistical model. Hereby we binarize the outcome variable DTG resistance, comparing DTG susceptible with any DTG resistance.

Sensitivity analysis S3: Risk factor analysis in the subset with RT sequences. As a considerable number of genotypic resistance tests in the main risk factor analysis did not cover the RT region, we considered the additional level `no RT sequenced`. As both for NRTI and NNRTI, genotypic resistance is predicted from RT, the `no RT sequenced` level resulted in collinearity. In S3, we confirm the findings in the subset of study participants where RT sequence was available. Additionally, we investigate impact of NNRTI resistance on DTG resistance.

Sensitivity analysis S4: Risk factor analysis using 3TC and TDF for calculating NRTI resistance. In light of the global roll-out of TLD (tenofovir lamivudine dolutegravir), we calculate in S4 the resistance levels based on resistance scores for 3TC and TDF only, and consider the maximum score of the two NRTIs to derive NRTI resistance level.

Sensitivity analysis S5: Risk factor analysis in the subset on a DTG regimen with two NRTI backbones. In S5, we confirm our findings in the subset of participants on a triple regimen comprised of DTG plus two NRTIs. This corresponds to 3TC, ABC, DTG (N=243, 65·7%); DTG, FTC, TDF (N=88, 23·8%); DTG, FTC, TAF (N=34, 9·2%); 3TC, AZT, DTG (N=3, 0·8%); 3TC, DTG, TDF (N=1, 0·3%), and ABC, DTG, TDF (N=1, 0·3%). We calculate NRTI resistance based on only the two NRTIs comprising the ART regimen of the study participants.

Sensitivity analysis S6: Using prior NRTI resistance as covariable in the risk factor analysis. The association of NRTI resistance with DTG resistance found in the risk factor analysis does not allow to determine which came first. In S6, we restrict the study population to those where genotypic resistance tests from earlier timepoints were available. We calculate prior NRTI resistance for earlier timepoints, taking the maximum in case multiple earlier genotypic resistance tests were available.

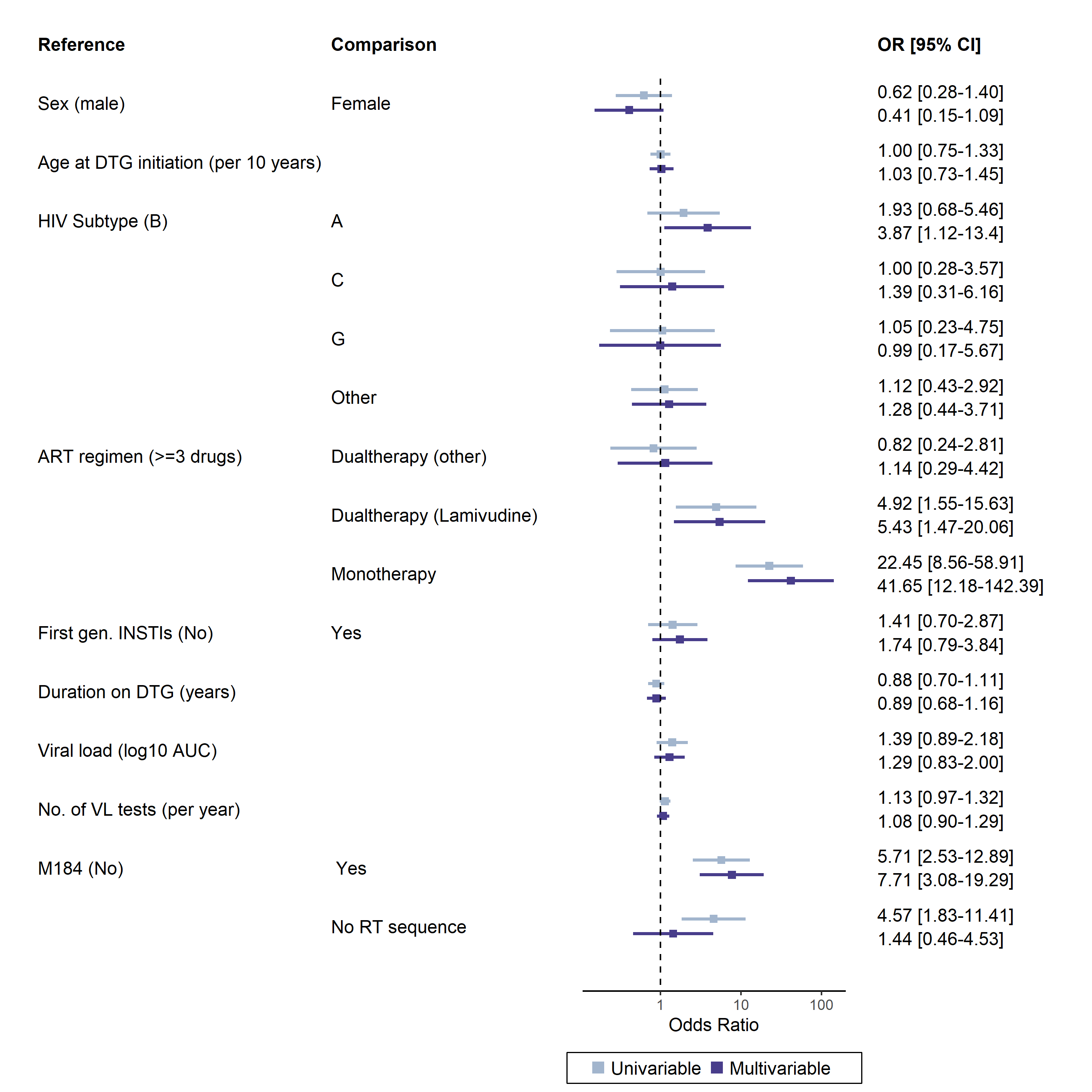

Supplemental figure 3: **Sensitivity analysis S1: Using M184I/V as covariable in the risk factor analysis**. Risk factors were assessed in the same population as in the main risk factor analysis. Cohorts were included as random effect.

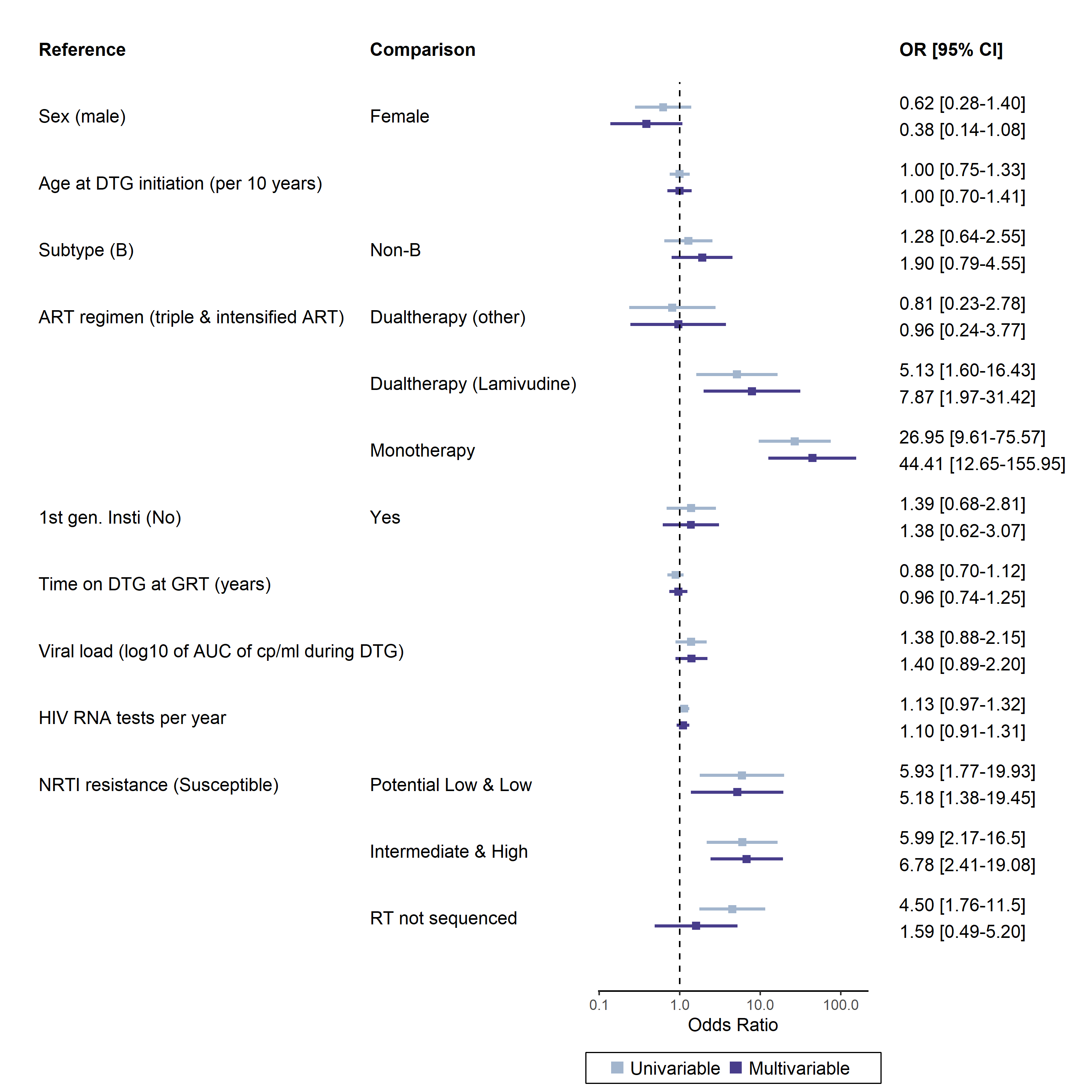

Supplemental figure 4: **Sensitivity analysis S2.** **Logistic regression for DTG resistance levels susceptibly vs. any DTG resistance**. Risk factors using logistic regression were assessed in the same population as in the main analysis. Cohorts were included as random effect.

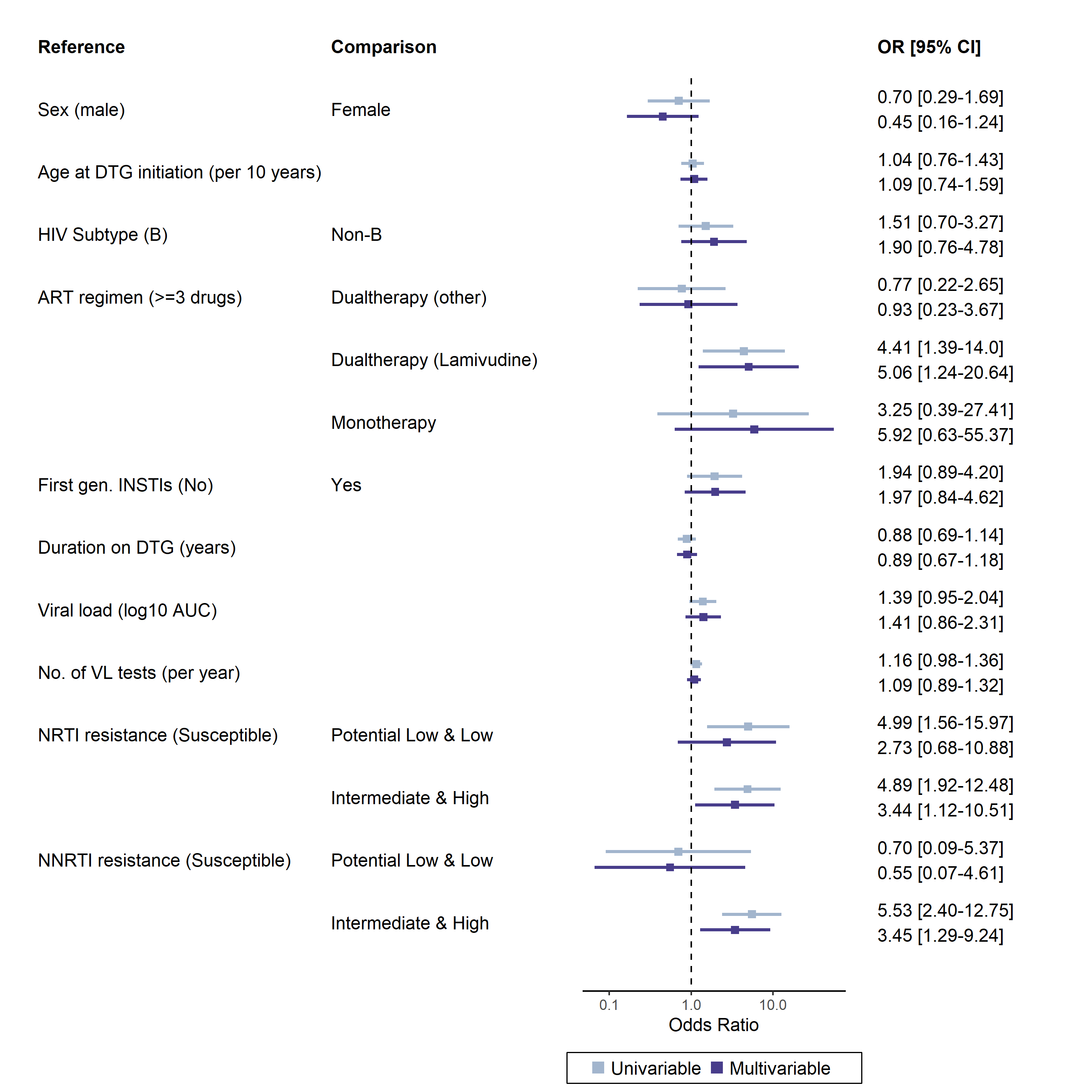

Supplemental figure 5: **Sensitivity analysis S3. Full RT sequence sensitivity analysis.** Risk factor analysis in the 613 out of 677 (90·5%) people considered in the main risk factor analysis where RT sequence was available. Participants where no RT sequence was available were excluded in this analysis, thus allowing to include NRTI and NNRTI resistance concurrently by resolving the collinearity of RT sequence availability.

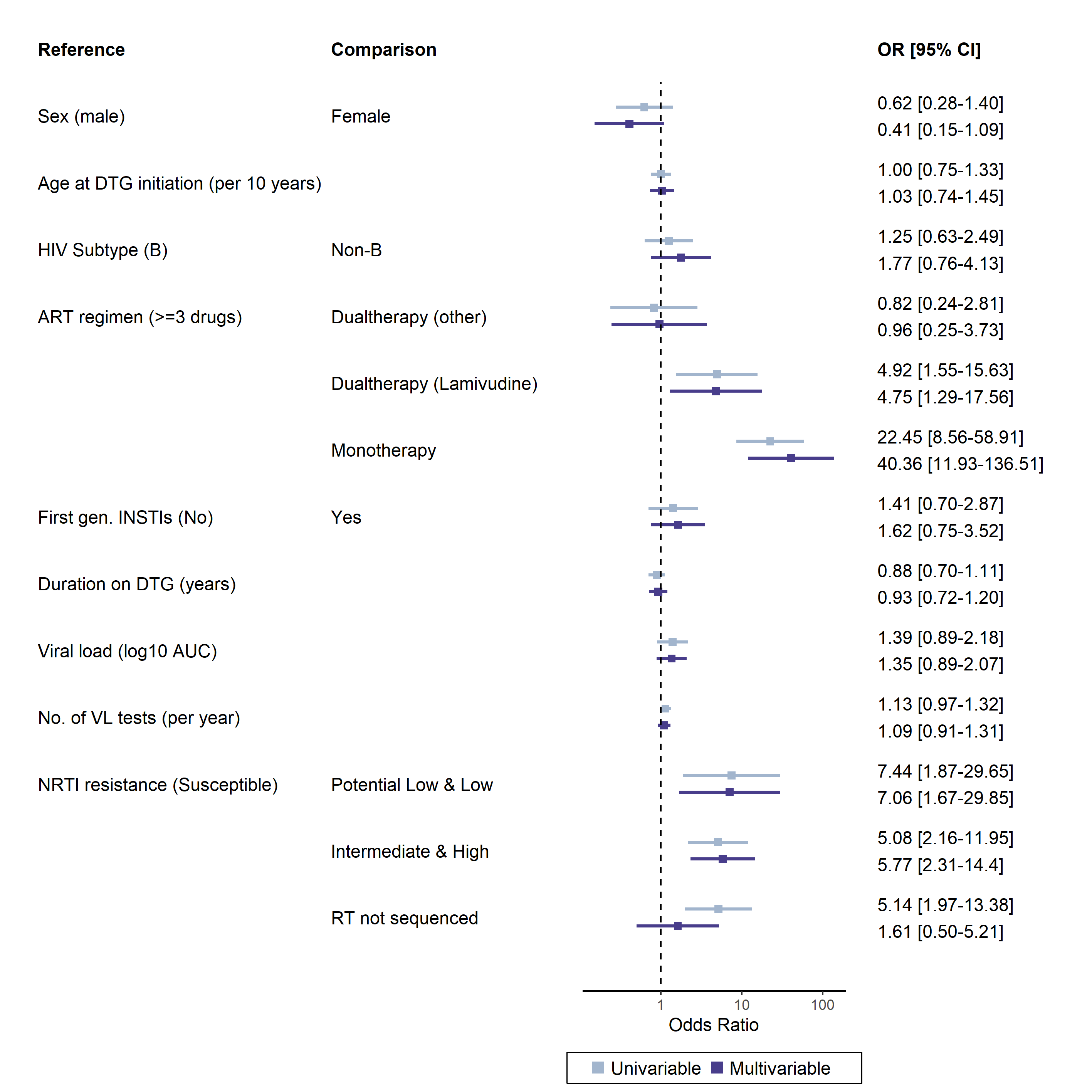

Supplemental figure 6 **Sensitivity analysis S4. NRTI resistance calculated based on 3TC and TDF**. Considering the wide use of TLD, we considered 3TC and TDF for calculating NRTI. Risk factors were assessed in the same population as in the main risk factor analysis. Cohorts were included as random effect.

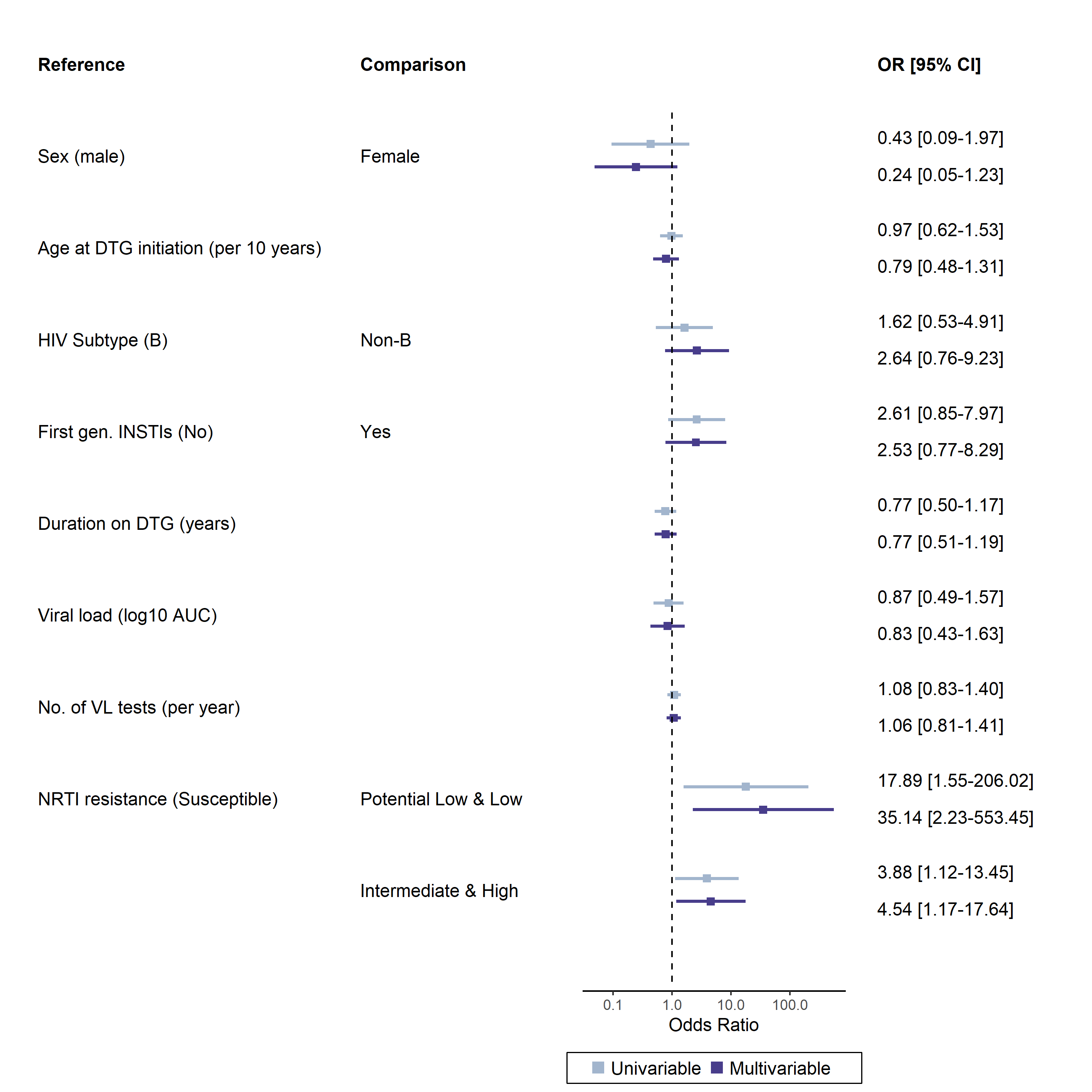

Supplemental figure 7: **Sensitivity analysis S5. Study participants on a triple regimen with DTG and two NRTIs**. NRTI resistance was calculated as maximum level to backbone drugs for each individual. ART regimen were as follows: 3TC, ABC, DTG (N=243, 65·7%); DTG, FTC, TDF (N=88, 23·8%); DTG, FTC, TAF (N=34, 9·2%); 3TC, AZT, DTG (N=3, 0·8%); 3TC, DTG, TDF (N=1, 0·3%), and ABC, DTG, TDF (N=1, 0·3%).

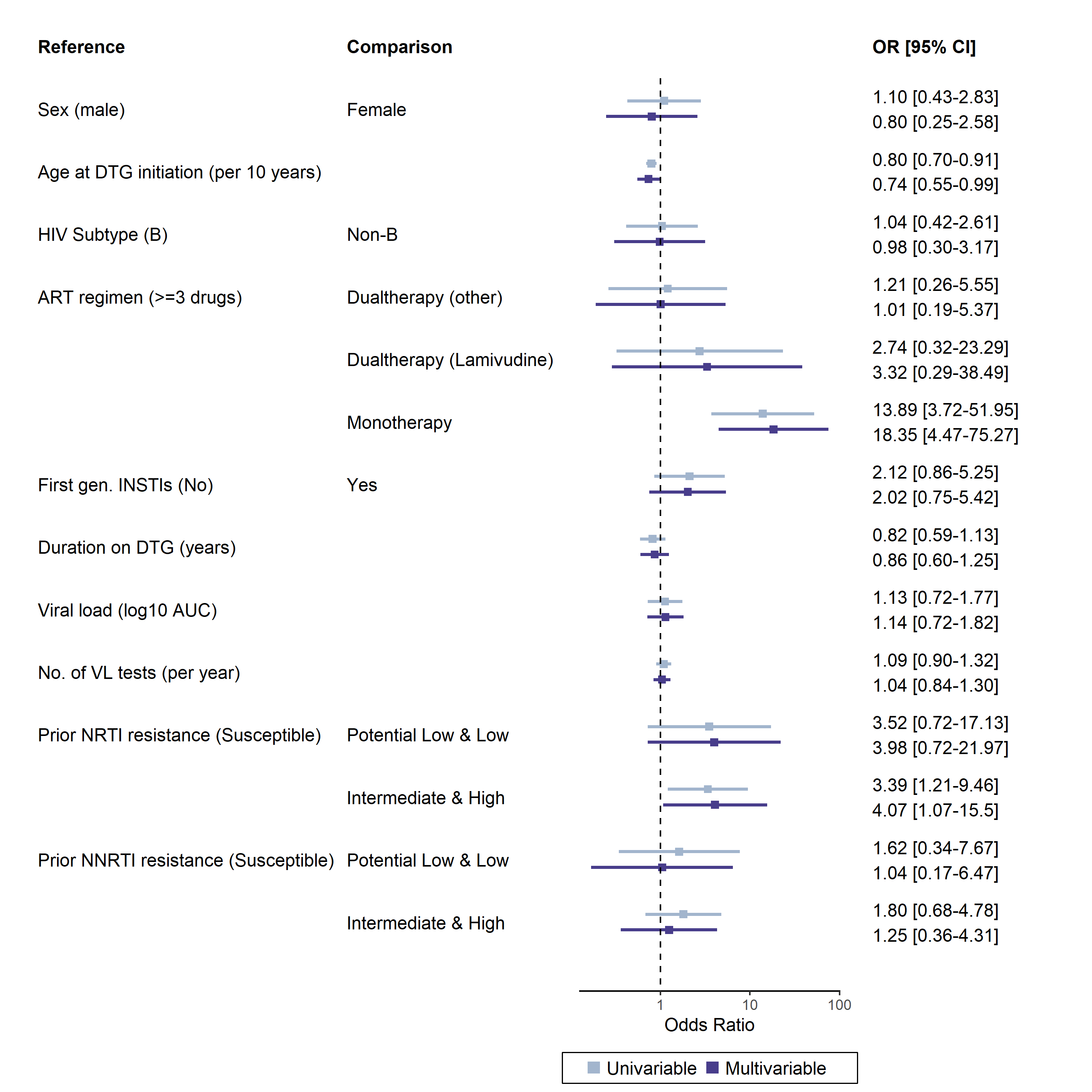

Supplemental figure 8: **Sensitivity analysis S6. Risk factor analysis including prior genotypic resistance tests.** For the 389 out of 677 people who had additional prior GRTs, NRTI and NNRTI resistance was calculated as in the main analysis for the prior resistance tests. We then considered the highest measured resistance level for NRTI and NNRTI, respectively, as prior resistance.
